# A Coupled Experimental and Statistical Approach for an Assessment of the Airborne Infection Risk in Event Locations

**DOI:** 10.1101/2022.01.10.22269028

**Authors:** Lukas Siebler, Torben Rathje, Maurizio Calandri, Konstantinos Stergiaropoulos, Bernhard Richter, Manfred Nusseck, Claudia Spahn

## Abstract

Operators of event locations are particularly affected by a pandemic. Resulting restrictions may cause uneconomical business. With previous models, only an incomplete quantitative risk assessments is possible, whereby no suitable restrictions can be derived. Hence, a mathematical and statistical model has been developed in order to link measurement data of substance dispersion in rooms with epidemiological data like incidences, reproduction numbers, vaccination rates and test qualities. This allows a first time overall assessment of airborne infection risks in large event locations. In these venues displacement ventilation concepts are often implemented. In this case simplified theoretical assumptions fail for the prediction of relevant airflows for infection processes. Thus, with locally resolving trace gas measurements and specific data of infection processes, individual risks can be computed more detailed. Via inclusion of many measurement positions, an assessment of entire event locations is possible. Embedding the overall model in a flexible application, daily updated epidemiological data allow latest calculations of expected new infections and individual risks of single visitors for a certain event. With this model, an instrument has been created that can help policymakers and operators to take appropriate measures and to check restrictions for their effect.

## 1 Introduction

The pandemic of SARS-CoV-2 forced cultural institutions such as theatres and music halls to restrict or cancel their programs, without knowing the actual infection risk at their location. In order to identify this risk and take appropriate measures scientific evidences and risk estimation models for these venues are required.

Airborne transmission of viruses is a complex process involving emission, dispersion in the room, and inhaling [1]. There are many calculation models for an aerosol transmission and infection risk of SARS-CoV-2 in indoor environments [2–7]. However, these calculators often require idealised assumptions (e.g. ideal mixed ventilation), which lose accuracy with the size of the room. Therefore, they mainly consider small to medium room sizes.

In larger event locations, displacement ventilation concepts are often implemented, whose virus transmission towards neighbors is challenging to predict. Unobjectionable vertical buoyancy flows are superimposed by critical horizontal flows due to disturbance effects (e.g. cold walls and leaky doors). The estimation becomes even more critical in large and complex rooms, when relevant boundary conditions are unknown. [8] Moreover, these models neglect the access probabilities of infectious persons, which can be derived from specific epidemiological data. In summary, it has to be stated, that the required overall risk assessment model for large locations is still missing.

Within the framework of a project at the Stuttgart State Theatre, locally resolved trace gas measurements were carried out to calculate the airborne infection risk. Based on these investigations, a coupled experimental and statistical model is essential.

Depending on the epidemiological data (incidence and reproduction number, vaccination rate and test quality) the access probability of an infectious person to an event location varies. Furthermore vaccinations prevent infections when sitting near an infectious person. All the data are highly relevant for an overall risk assessment, especially for locations with several hundred people. Hence, the research objective is to develop a coupled experimental and statistical model that accounts for these data. The results will provide scientifically based recommendations for actions non-pharmaceutical interventions (NPI) to maintain the operation of cultural institutions under pandemic conditions.

## 2 Model for an overall assessment of infection risks

The approach of the model can be separated in several sections, which are interacting with each other. This calculation procedure is shown in figure 1 and assists to comprehend the structure of the paper by means of a section assignment.

**Fig 1.**
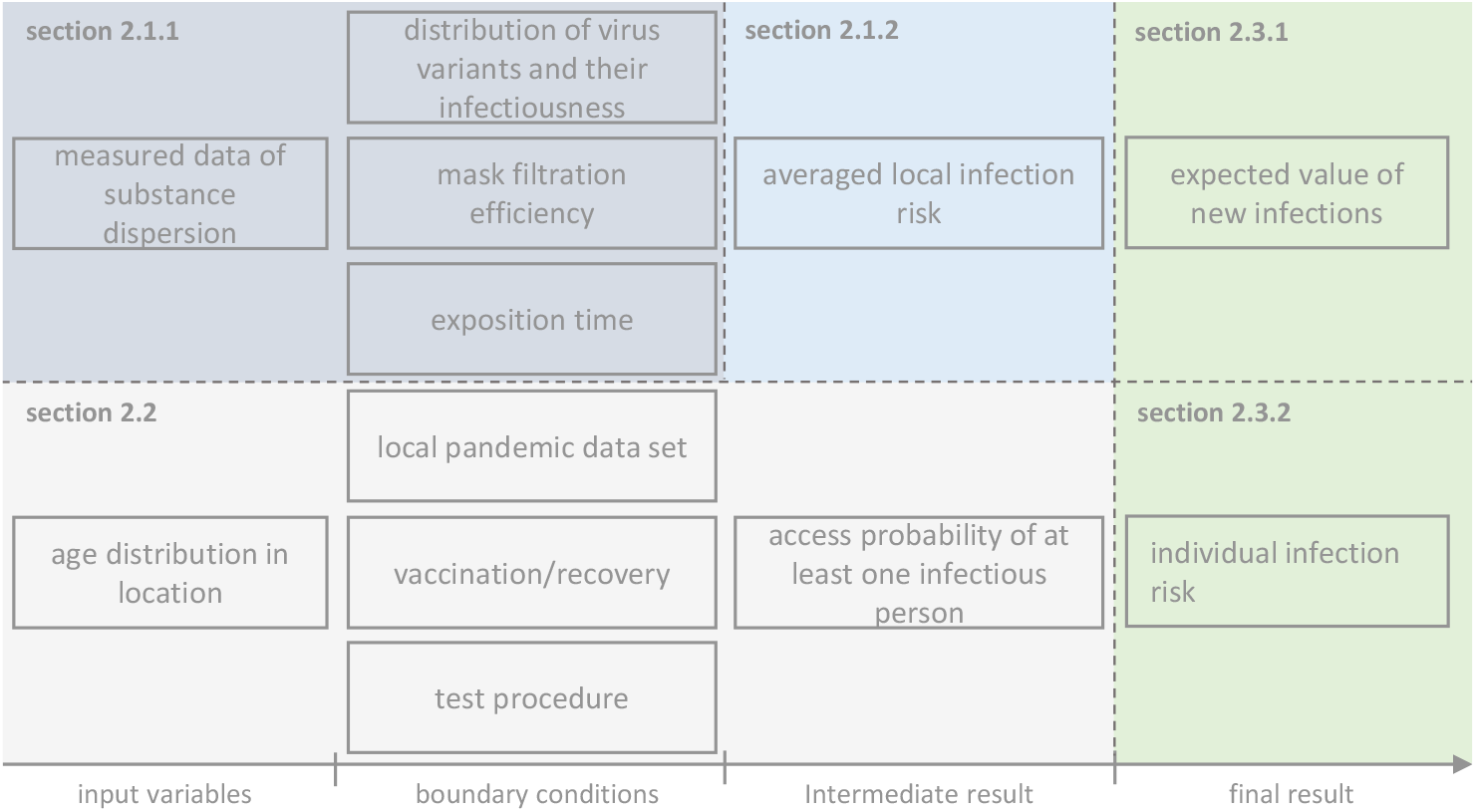
structure of the combined model for an overall assessment of infection risks

Using measured data of substance dispersion in combination with certain boundary conditions allows a tranfer to an estimated airborne infection risk (see section 2.1.1). Due to the combination of many individual results an averaged local infection risk (see section 2.1.2) can be calculated. The access probability of an infectious person is calculated based on the age distribution in location and epidemiological data (see section 2.2). In section 2.3.1 the expected value of new infections as a final result is calculated. In order to compute an individual infection risk as a second result the individual vaccination/recovery status is required (see section 2.3.2). In the following, each model section is presented separately.

### 2.1 Measurements

The focus of previous indoor air investigations using trace gas, also conducted at the University of Stuttgart, has mostly been on evaluating the ventilation effectiveness rather than infection risks [9]. For this new purpose, however, such measurements are also well suited, whereby a transfer to viral loads has to be performed. The method even provides reliable results for unknown air flows resulting from partly uncertain boundary conditions.

#### 2.1.1 Substance dispersion measurements at individual positions

For transferring measurement data into airborne infection risks a mathematical approach is required. The model of *Wells et al*. and *Riley et al*. [10, 11] allows an estimation of the predicted infection risk via aerosols (PIRA). They introduced quanta as a fictitious unit for an amount of inhaled viruses which lead to an infection with a certain probability. As a result the following correlation is announced:

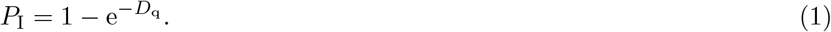

with *P*_I_ and *D*_q_ as PIRA and dose of inhaled quanta, respectively. *Siebler et al*. [8] suggests an experimental approach with trace gas for ventilation with outdoor air exchange. It is based on releasing a certain rate of gas with a mass flow controller. Measuring the neighboring concentrations with a gas analyser enables users the quantification of substance dispersion in general. Assuming that trace gases are dispersed in the same way as relevant virus-bearing aerosols [12], a transfer to infection risk is possible. In order to link trace gas to quanta and to account for mask filtration effects the following approach is introduced:

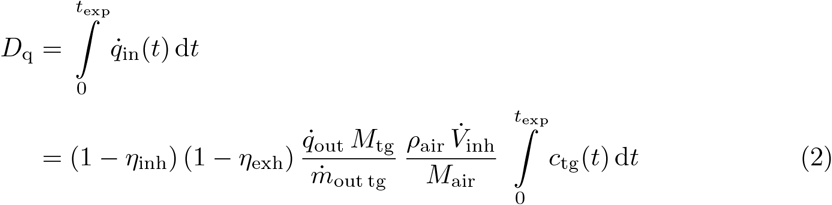

with 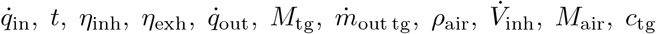 as inhaled quanta rate, time, mask filtration efficiency for inhaling/exhaling, exhaled quanta rate, molar mass of trace gas, mass flow of trace gas (output), density of air, inhalation volume flow, molar mass of air and measured trace gas concentration respectively. In order to calculate the dose a numerical integration of trace gas concentration is needed. Filling in equation (1) results in the predicted infection risk via aerosols for a certain position. [8]

#### 2.1.2 Transfer of individual measurement positions on entire location

Quanta concentrations of neighbours sitting near an infectious person vary depending on disturbance influences of the ventilation concept. Particularly displacement ventilation systems are sensitive for disturbance effects. If measurement positions are limited due to the equipment these can be quantified by random sampling. For more conservative consideration critical assumed areas (e.g. dead water areas and low ceiling heights) shall predominate. Therefore, transfer methods are needed, of which one potential method is presented.

First the measurement positions in the neighborhood of an infectious person should be well distributed. A 360° circumferential arrangement of measurement positions is recommended because critical horizontal air flows are not known and might predominate over the momentum of exhalation. In figure 2 each neighbourhood is divided into rings – in that case one for close (black line) and one for far neighbours (dark blue line). Besides this division concept there are a few alternatives. Representative measurement results included in these rings are averaged. Finally all neighbourhoods are averaged again to have an overall assessment for a statistically prediction of an arbitrary place. This procedure is outlined in figure 2 for an exemplary occupancy density of 50 %. For any other density an alternative positioning might be more suitable.

**Fig 2.**
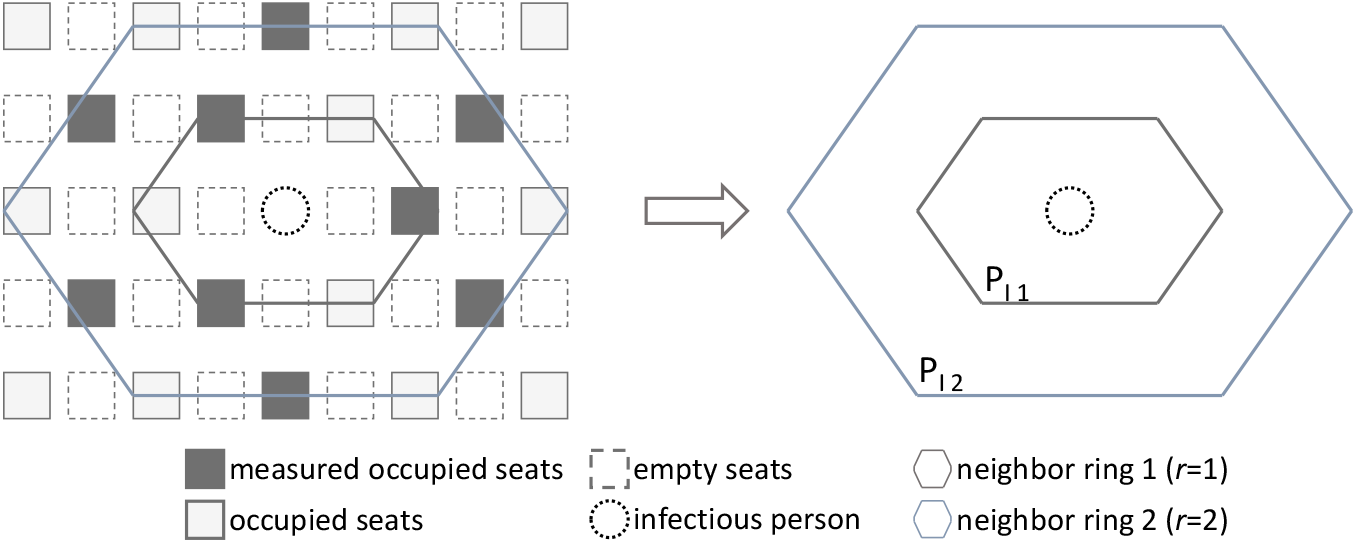
procedure of averaging measurement data for an overall PIRA

The expected value of new infections in several neighbour-rings of one available infected person can be calculated generally as:

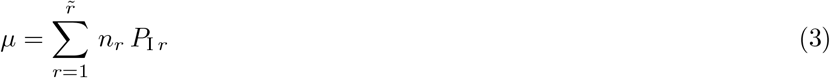

With 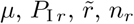 as expected value of new infections, infection risk in the certain neighbor ring *r*, number of considered neighbor rings and number of persons in the certain neighbor ring *r* respectively.

### 2.2 Access probability of one infectious person

The probability *P*_acc_ of infectious persons in an event location can be calculated with the binomial distribution of Gauss [13], if the probability *p*_acc_ of an individual infectious person gaining access is known. Equation 4 shows the calculation in detail:

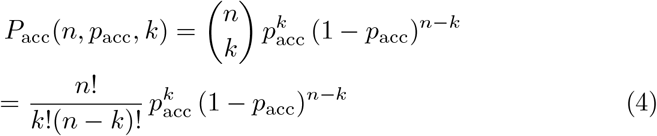

with *n* and *k* as number of persons in the location and number of infectious persons respectively.

In order to keep the complexity and the effort of the statistical model in acceptable range it is only practical for maximum one infectious person in the location. For small *p* (for high vaccination rates and a high quality test procedure) the following assumption and resulting equation is appropriate and above all conservative [13]:

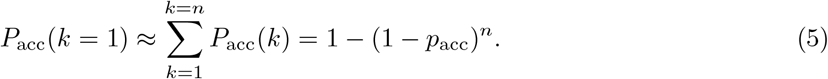

For an accurate the identification of *p*_acc_, different types and dimensions of classifications are possible in principle, depending on the accessibility of the data. In this case, the status of vaccination is significant. A subdivision in not vaccinated, recovered and vaccinated allows higher accuracy for predicting *p*_acc_. An additional subdivision in several different vaccine types due to different effectiveness is specifying the results further.

For event locations, the age distribution of visitors might be a relevant parameter. The vaccination rate and the vaccine types are often age dependent. In table 2, one exemplary classification is shown.

**Table 1.** Nomenclature

| quantity | description | unit |
| --- | --- | --- |
| **Acronyms** |  |  |
| FFP2 | filtering facepiece (class 2) |  |
| SARS-CoV-2 | Severe acute respiratory syndrome coronavirus 2 |  |
| mRNA | messenger ribonucleic acid |  |
| NPI | non-pharmaceutical interventions (NPI) |  |
| PCR | polymerase chain reaction |  |
| PIRA | predicted infection risk via aerosols |  |
| **Roman Symbols** |  |  |
| $c$ | concentration | $\text{mol mol}^{-1}$ |
| $D$ | dose | — |
| $I$ | incidence number | — |
| $k$ | number of infectious persons | — |
| $M$ | molar mass | $\text{kg mol}^{-1}$ |
| $n$ | number of persons | — |
| $\dot{m}$ | mass flow | $\text{kg h}^{-1}$ |
| $P$ | overall probability for $k$ infectious persons in the location | — |
| $p$ | probability of one single person being infected | — |
| PIRA | predicted infection risk via aerosols | % |
| $R$ | reproduction number | — |
| $q$ | quanta rate | $\text{h}^{-1}$ |
| $t$ | time | h |
| $\dot{V}$ | volume flow | $\text{m}^3 \text{h}^{-1}$ |
| **Greek Symbols** |  |  |
| $\eta$ | efficiency | — |
| $\mu$ | expected value of new infections | — |
| $\rho$ | density | $\text{kg m}^{-3}$ |
| $\tau$ | infection transmission constant | $\text{d}^{-1}$ |
| **Subscripts** |  |  |
| 0 | initial state |  |
| air | air |  |
| acc | access |  |
| crit | critical |  |
| exh | exhalation |  |
| exp | exposition |  |
| i | vaccination / recovery status class |  |
| I | infection risk via aerosols |  |
| in | in |  |
| ind | individual |  |
| inf | infection |  |
| inh | inhalation |  |
| j | age class |  |
| out | out |  |
| q | quanta |  |
| r | neighbor ring |  |
| tg | trace gas |  |
| total | total |  |
| fnt | false negative test |  |
| ∼ | number of classes | |

**Table 2.**
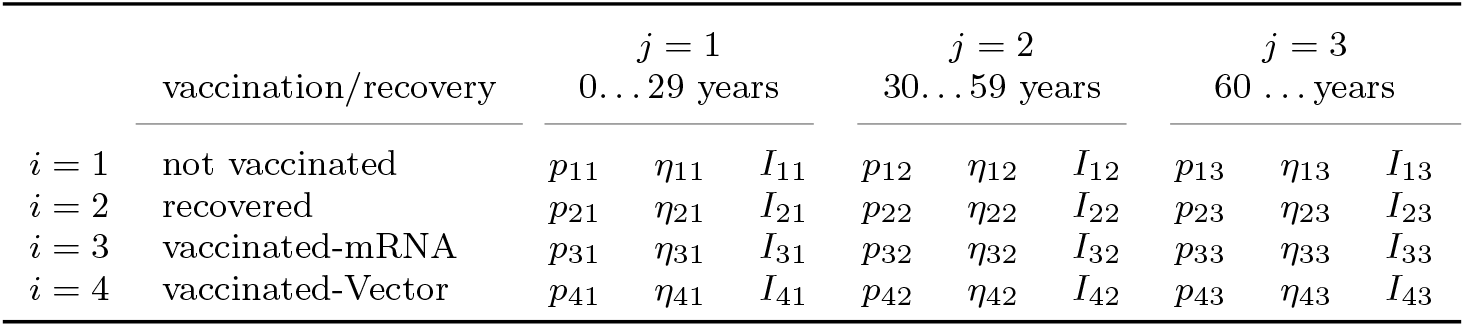
classification of visitors and associated data

For each class combination there is a probability of *p*_*ij*_ for a person belonging to it. For instance, an arbitrary person of possible visitors, has the probability *p*_31_ to be mRNA-vaccinated and between 0… 29 years old with a related vaccine effectiveness *η*_31_ and incidence number *I*_31_.

Incidence numbers in general describe the rate of (new) infections per time and inhabitants. It is assumed that once someone knows that they are infectious they generally stay at quarantine or at least they do not try to get access to an event location. Therefore, predicting incidence rates is relevant to the probability of gaining access being infectious *p*_acc ij_ because presymptomatic, asymptomatic, and unwittingly infectious persons are critical.

This forecast can be estimated by the reproduction number *R* (R-value), the mean duration between being infected and infecting the next person Δ*t*_inf_ and the mean critical duration between one becomes infectious and the knowledge about it Δ*t*_crit_. *R* describes how many persons one infectious person infects in average. For even higher accuracies, reproduction numbers could optionally be further classified by age groups and vaccination/recovery status. The following equation shows the correlation between the aforementioned values assuming an exponential course:

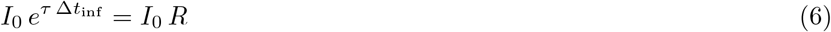

with *I*_0_, *τ*, Δ*t*_inf_, *R* as initial overall incidence (no age and vaccination/recovery status related separation), infection transmission constant in 1/d, mean duration between being infected and infecting the next person and reproduction number (R-value) respectively.

The course of the incidence value correlates with the constant *τ*, which can be derived from equation 6:

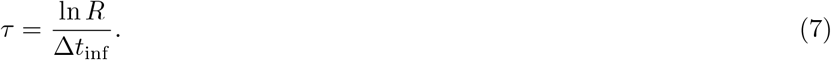

To estimate the critical ratio of infectious persons, who do not know about their infectiousness at a certain day, it is useful to predict the incidence course up to the duration Δ*t*_crit_ later. The integral of that course over duration describe persons, who could be infectious over that period (Δ*t*_crit_) without knowing it. It is calculated as:

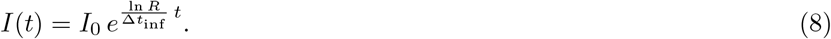

The access probability of an infectious person of a certain class combination (e.g. 23) is depending on its certain incidence (e.g. *I*_23_). In addition, the test quality with regard to false negative results of different principles (antigen, polymerase chain reaction (PCR)) is considered [14] [15].

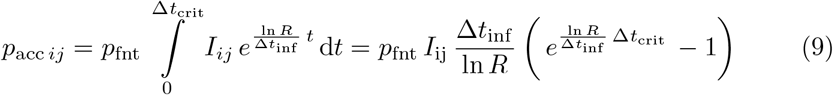

with *p*_acc *ij*_, *I*_*ij*_, *p*_fnt_ as access probability of an infectious person of a certain class combination *ij*, incidence of a certain class combination *ij* and probability of a false negative test result (if no test is done *p*_fnt_ = 1) respectively.

The calculation process of an overall probability for one arbitrary person being infectious and getting access is shown in figure (3). It is worth mentioning that the classifications can be extended as desired.

**Fig 3.**
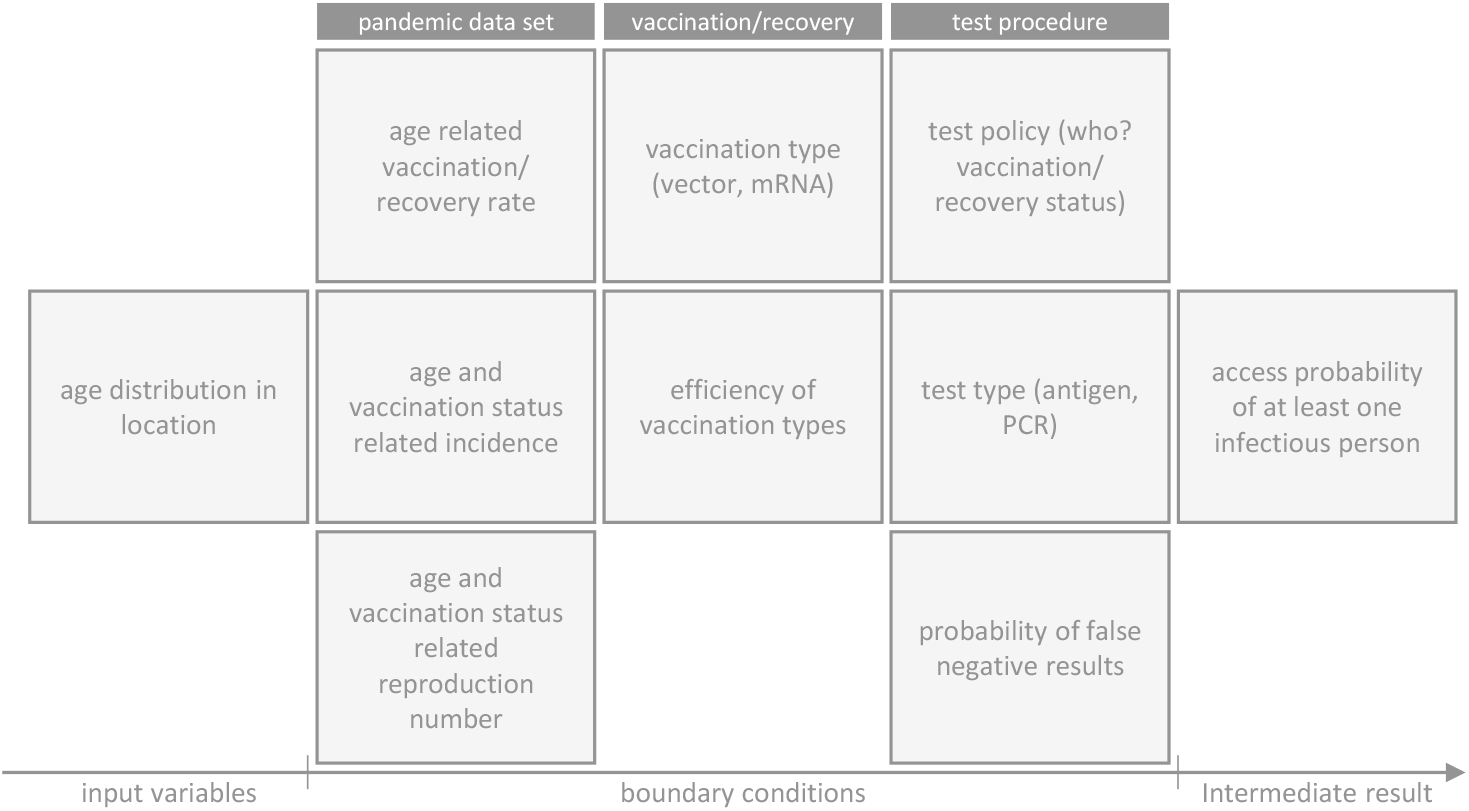
calculation process for an access probability of at least one infectious person

The process of the shown scheme can be calculated as sums (number of sums equal to the dimension of the classification) of the proportion of each class combination *p*_*ij*_ and its certain probability of an infectious person inside *p*_acc *ij*_ :

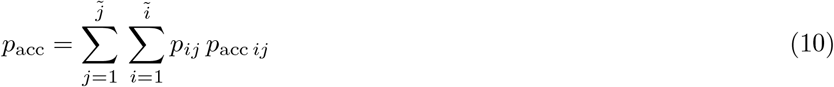

Equations (9) and (10) filled in (5) result for the access probability of an infectious person in:

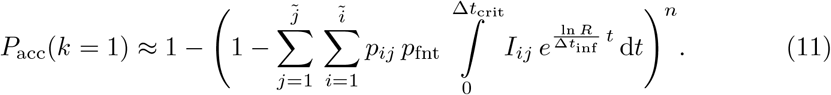

### 2.3 Model output

In this section the two outputs of the model are derived. Besides the expected value of new infections, which is relevant for operators and policy makers, an infection risk for an individual person is determined.

#### 2.3.1 Expected value of new infections

Once an infectious person is in the location, the vaccination status of their neighbors is relevant. Taking the vaccination impact twice into account (access probability and neighbor vaccination status) is noteworthy for this model. Due to the classification, see table 2, equation (3) becomes more complex. Thereby the probability of neighbors belonging to a certain class combination is taken into account. In the following equation the union between measurement and statistical data occurs.

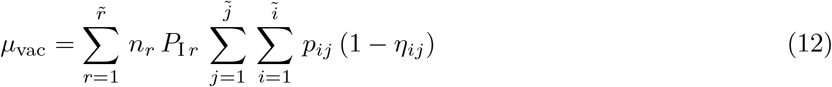

with *µ*_vac_, *η*_*ij*_ as expected value of new infections regarding the vaccination status and their efficiency respectively.

Combining equation (12) with the probability of access (equation (11)) results in the overall expected value of new infections:

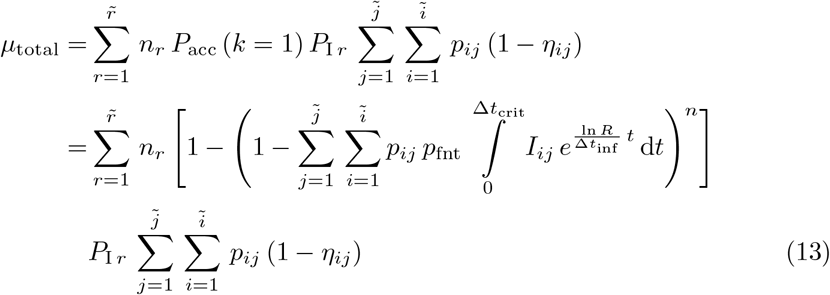

with *µ*_total_ as expected value of new infections regarding entrance probability and vaccination status. This calculation provides a valuable instrument for operators of event locations and politicians for a total risk assessment of infections.

#### 2.3.2 Infection risk for an individual person

With regard to an individual person the probability of being exposed in a certain neighbor ring of an infectious person has to be taken into account as follows:

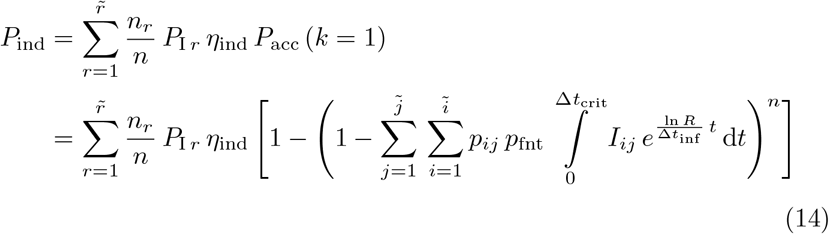

with *P*_ind_, *η*_ind_ as individual infection risk of one certain person and event date and efficiency of the individual vaccination status respectively. This calculation enables visitors to estimate their individual infection risk regarding the decision of participating an event.

#### 2.3.3 Limitations and boundaries of the model

The coupled experimental and statistical model assumes that maximum one infectious person is present. Due to superposition of trace gas results risk assessments for more than one infectious person could be calculated. However, the effort for this statistical model adjustment would become laborious.

Figure 4 shows the deviation between 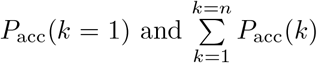 (which are assumed to be approximately equal (equation (5)) in this study) for *n* = 100 and *n* = 500 visitors. For *n* = 500 up to *p*_acc_ *≈* 1 *×* 10^4^ and *n* = 100 up to *p*_acc_ *≈* 1 *×* 10^3^ the deviations are assumed low. Hence even for large locations the model stays conservative and also acceptable for small *p*_acc_. With this approach users could estimate whether this model is still accurate enough. Otherwise, operation with such boundary conditions is questionable anyway.

**Fig 4.**
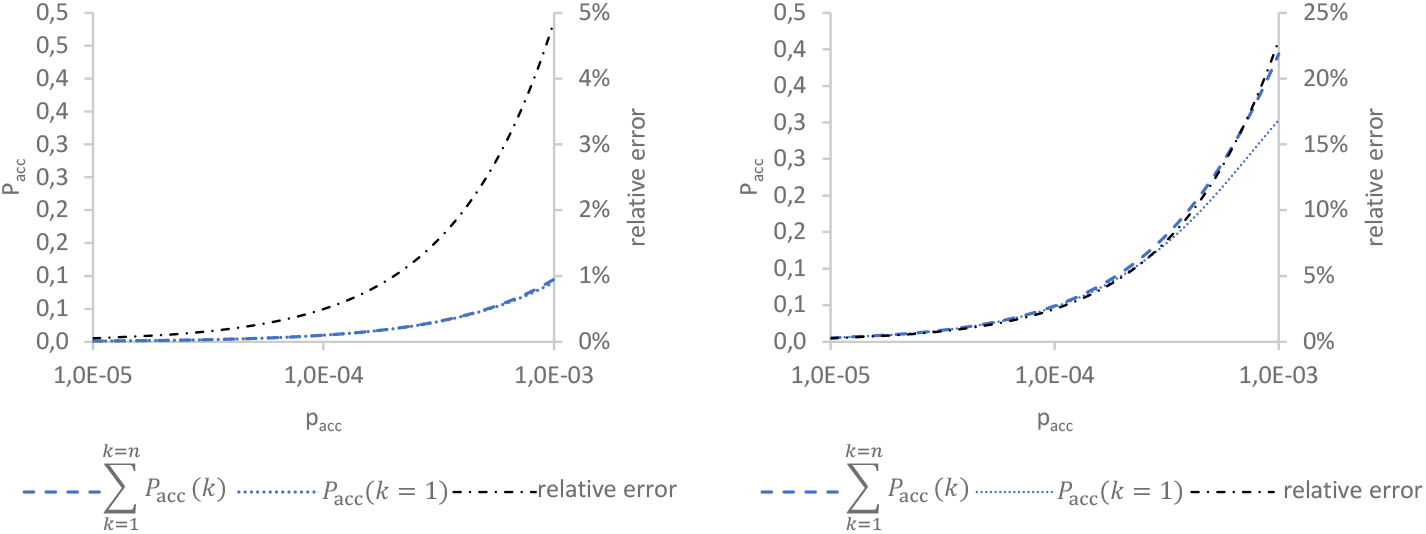
error estimation between 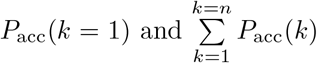, left *n* = 100, right *n* = 500

## 3 Discussion

In order to develop a robust mathematical and statistical model, there are several assumptions needed. One of these relates to the transfer of individual measurement positions to entire locations. It is supposed that the number of seats each ring contains is constant. However, it varies at edge positions of an infectious person. This assumption reduces the accuracy of the model marginally, but it is still conservative.

Besides, the structure of the coupled model is adapted to visitors instead of artists and staff. To analyse the air flow interactions between audience and stage further trace gas measurements should be carried out.

It should be noted that included statistical data must be robust and detailed. Given the fact, that infection processes are dynamic, possible virus mutations should be considered when calculating the infection risk. It is assumed that responsible action will be taken and that quarantine measures will be followed in the case of known infection (no attendance at events). Otherwise, the accuracy of the model will be affected. Furthermore, in order to validate the model appearing infections can be taken into account (e.g. *p*_fnt_ or the classification in general).

## 4 Conclusion

This paper presents a coupled experimental and statistical approach for an assessment of the airborne infection risk in event locations. Combining data of trace gas measurements and statistical data in one model provides two valuable results: For operators and policy makers it delivers an expected value of new infections for a certain event. For society an individual infection risk for this event is given, which might help in deciding whether to participate in certain events. Both results are valuable tools that can make it possible to maintain social life despite an ongoing pandemic. Hence, the objective of improving the validity of the existing models described in section 1 was achieved by developing this coupled experimental and statistical model.

## Data Availability

All data produced in the present study are available upon reasonable request to the authors

## Acknowledgments

We would like to thank Dr. Tjibbe Donker who supported us with his expertise regarding the infection process. Another special thanks goes to Mr. Donagh Hennessy for his scientific editing services.

## Declarations

### Data availability statement

Datasets generated/analysed during the current study are available from the corresponding author on reasonable request.

### Ethics approval statement

Not applicable.

### Patient consent statement

Not applicable.

### Permission to reproduce material from other sources

Not applicable.

### Authors’ contributions

LS: Conceptualization, methodology and writing original draft. TR: Participation in the conception and writing— review & editing. MC: Writing— review & editing and visualization. KS: Writing— review & editing. MN: Conceptualization, writing— review & editing. BR: Writing— review & editing. and CS: Supervision writing— review & editing

